# On the ability to extract MLVA profiles of *Vibrio cholerae* isolates from WGS data generated with Oxford Nanopore Technologies

**DOI:** 10.1101/2023.02.17.23286076

**Authors:** Jérôme Ambroise, Bertrand Bearzatto, Jean-Francois Durant, Leonid M. Irenge, Jean-Luc Gala

**Author notes:** Corresponding author Jérôme Ambroise.

## Abstract

Multiple-Locus Variable Number of Tandem Repeats (VNTR) Analysis (MLVA) is widely used by laboratory-based surveillance networks to subtype pathogens causing foodborne and water-borne disease outbreaks. The *MLVAType* shiny application was previously designed to extract MLVA profiles of *Vibrio cholerae* isolates from WGS data, and provide backward compatibility with traditional MLVA typing methods. The previous development and validation work was done on short (pair-end 300 and 150 nt long) reads from Illumina MiSeq and Hiseq sequencing. In the initial phase of this work, the *MLVAType* application was validated on long reads generated by Oxford Nanopore Technologies (ONT) sequencing platforms. The MLVA profiles of *V. cholerae* isolates (n=9) from the Democratic Republic of the Congo were produced using the *MLVAType* application on WGS data. The WGS-derived MLVA profiles were extracted from canu (v.2.2) assemblies obtained through MinION and GridION sequencing by ONT. The results were compared to those obtained from SPAdes assemblies (v3.13.0; k-mer 175) generated from short-read (pair-end 300-bp) data obtained by MiSeq sequencing, Illumina, taken as a reference. For each isolate, the MLVA profiles were concordant for all three sequencing methods, demonstrating that the *MLVAType* application can accurately predict the MLVA profiles from assembled genomes generated with long-reads ONT sequencers.

In the final phase of this study, we conducted phylogenomic analysis on data generated by both sequencing technologies, highlighting the superior resolution of Illumina short-read sequencing compared to the ONT-based approach. However, there was a remarkable concordance between isolate clusters identified using ONT-based MLVA profiles and those derived from the short-read-based phylogenomic analysis. This striking agreement enabled us to identify specific benefits and drawbacks of both technologies.

## Background

Multiple-Locus Variable Number of Tandem Repeats (VNTR) Analysis (MLVA) is widely used by laboratory-based surveillance networks for subtyping pathogens causing foodborne and water-borne disease outbreaks. We recently demonstrated that WGS data generated with short-read Illumina sequencing technology can be used to extract MLVA profiles of *V. cholerae* isolates from WGS data while maintaining backward compatibility with traditional MLVA typing methods (1). The percentage of censored estimations in MLVA profiles generated from WGS data was inversely proportional to the k-mer parameter used during genome assembly. However, preventing censored estimation was possible by using a longer k-mer size (e.g. 175) even though the original SPAdes v.3.13.0 (2) software did not propose this k-mer size.

Both MinION and GridION ONT sequencers are quickly gaining popularity because the long sequence reads enable to assemble contiguous microbial genome. However, their base-calling accuracy is significantly lower than that obtained with Illumina short reads, although the resolution of this shortcoming is steadily improving. More specifically, it is well known that ONT sequencers have difficulty in accurately sequencing low-complexity regions, such as homopolymers (3).

The current study was carried out in two stages: The goal of the first phase was to broaden the scope of application of our previous *MLVAtype* shiny application by comparing and validating MLVA results obtained when using WGS data obtained with MinION and GridION sequencing. Accordingly, we compared the MLVA profiles obtained using the three methods on a series of *V. cholerae* strains.

In the final phase, we conducted a comparative analysis to assess the advantages and disadvantages of short-read and long-read sequencing technologies for determining the phylogenetic relationships between strains. Assembled genomes derived from both Illumina and ONT sequencing were analysed using k-SNP. Subsequently, the resulting phylogenomic trees were compared.

## Method

### (a) Sample collection and sequencing technology

For the first phase of the study, nine *V. cholerae* isolates were selected from a collection of isolates characterised in a recent study conducted in the DRC between 2014 and 2017 (4).

Whole genome sequencing of selected *V. cholerae* isolates was performed using two technologies: Illumina (MiSeq and NextSeq 100) and ONT (MinION and GridION). Regarding Illumina technology, sequencing libraries were prepared starting from 70 ng of genomic DNA from DRC isolates according to Illumina DNA Prep protocol (Illumina, San Diego, CA, USA). Briefly genomic DNA were simultaneously fragmented and tagged with sequencing primers in a single step using Bead linked transposomes. Tagged DNAs were then amplified with a 12-cycle polymerase chain reaction (PCR) to generate sequencing ready DNA fragments by adding indexes and adapters. Final libraries were then cleaned up with AMPure beads, and subsequently loaded on a MiSeq for a paired-end 2 x 300 nt sequencing run using MiSeq Reagent kit V3 (600 cycles) (Illumina, San Diego, CA, USA).

ONT Long reads libraries were generated starting from 400 ng of high molecular weight genomic DNA (GQN > 8). DNA was firstly fragmented to an average fragment length of 11,6 kb using Covaris g-TUBES (Covaris, Woburn, WA, USA). Libraries were then prepared and barcoded according to the Ligation Sequencing genomic DNA – DNA Barcoding kit SQK-NBD112.24 protocol from ONT. The nine libraries generated were multiplexed and loaded on 2 FLO-MIN112 (R10 version) flow cells. Sequencing was undertaken on a MinION Mk1C and a GridION for 72 hours.

For the second phase of the study, we selected 32 additional V cholerae samples from our isolate collection (5). During this phase, both ONT and Illumina sequencing were carried out with the following modifications: for Illumina sequencing, libraries were prepared using the same Illumina DNA Prep protocol. We adapted the double-sided bead purification procedure to purify libraries with an average size of (450 bp), which is compatible with paired-end (2x150 pb) sequencing on a NextSeq 1000 sequencer.

For ONT sequencing, libraries were constructed using 100 ng of high molecular weight genomic DNA and the Rapid sequencing DNA V14-Barcoding kit (SQK-RBK114.96) protocol. These libraries were then sequenced for 72 hours on a GridION using R10.4.1 Flow cell.

### (b) WGS assembly and MLV profiling

WGS data generated with Illumina MiSeq were assembled into contigs using SPAdes v.3.13.0 (2) and a k-mer value of 175. Considering that this k-mer value is longer than what is offered by this version of the software, editing the software source code was required. WGS data generated with ONT MinION and GridION were assembled into contigs using canu v.2.2 (6).

For each isolate and each sequencing platform, the MLVA profiles were extracted from the assembled contigs using the *MLVAtype* algorithm implemented in an R shiny application and freely available at https://ucl-irec-ctma.shinyapps.io/NGS-MLVA-TYPING/. This application enables the user to upload a list of draft genomes and the nucleotide sequences of the motifs. It was used to predict MLVA profiles associated to the *V. cholerae* loci listed in Table 1 as in our previous study.

**Table 1.**
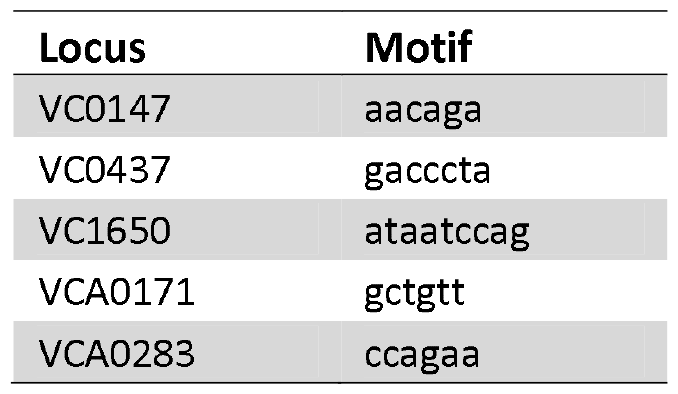
Loci and motifs characterising the MLVA profiles of *V. cholerae*

In the second phase of the study, we used k-SNP.3.0 (7) to analyse assembled genomes from short-reads (Illumina) and long-reads (ONT), and determine the phylogenomic relationships among *V. cholerae* strains. The MLVA profiles were also derived from long-reads derived genomes and analysed alongside with the phylogenomic trees.

## Results

Table 2 and Table 3 provide an overview of sequence quality reported for Illumina MiSeq, MinION and GridION, respectively. As expected, the forward Illumina MiSeq reads were of higher quality than the reverse reads.

**Table 2.** Quality control metrics of Illumina MiSeq reads

| Isolate | Number of reads | Length of reads | Positions in forward reads with median phred score >30 | Positions in reverse reads median phred score >30 |
| --- | --- | --- | --- | --- |
| CTMA-1426 | 2 x 2 228 605 | 2 x 300 | 0.983 | 0.751 |
| CTMA-1427 | 2 x 1 521 677 | 2 x 300 | 0.944 | 0.761 |
| CTMA-1432 | 2x 2 195 887 | 2 x 300 | 0.983 | 0.761 |
| CTMA-1435 | 2 x 1 613 586 | 2 x 300 | 0.9 | 0.754 |
| CTMA-1461 | 2 x 2 289 895 | 2 x 300 | 0.987 | 0.761 |
| CTMA-1473 | 2 x 2 525 965 | 2 x 300 | 0.987 | 0.761 |
| CTMA-1402 | 2 x 1 990 392 | 2 x 300 | 0.934 | 0.744 |
| CTMA-1421 | 2 x1 949 341 | 2 x 300 | 0.987 | 0.754 |
| CTMA-1424 | 2 x 1 955 144 | 2 x 300 | 0.924 | 0.764 |

**Table 3.** Quality control metrics of ONT MinION and GridION long reads

| ONT sequencing platform | Isolate | Number of reads | Length of reads min – max | median of reads length | Positions with median phred score >10 |
| --- | --- | --- | --- | --- | --- |
| MinION | CTMA-1426 | 190 186 | 69 - 63 195 | 1599 | 0.996 |
|  | CTMA-1427 | 87 929 | 70 - 43 611 | 1492 | 0.942 |
|  | CTMA-1432 | 112 216 | 68 - 61 372 | 759 | 0.985 |
|  | CTMA-1435 | 143 620 | 70 - 53 139 | 1444 | 0.990 |
|  | CTMA-1461 | 163 440 | 68 - 109 071 | 1612 | 0.978 |
|  | CTMA-1473 | 98 596 | 72 - 104 391 | 4162 | 0.989 |
|  | CTMA-1402 | 113 252 | 75 - 44 694 | 2098 | 0.989 |
|  | CTMA-1421 | 95 725 | 74 - 62 565 | 2604 | 0.991 |
|  | CTMA-1424 | 1 372 705 | 65 - 127 859 | 624 | 0.999 |
| GridION | CTMA-1426 | 122 597 | 67 - 43 560 | 1840 | 0.993 |
|  | CTMA-1427 | 57 920 | 70 - 54 700 | 1764 | 0.982 |
|  | CTMA-1432 | 71 818 | 73 - 43 924 | 857 | 0.995 |
|  | CTMA-1435 | 93 995 | 69 - 63 410 | 1683 | 0.991 |
|  | CTMA-1461 | 10 6175 | 69 - 53 447 | 1869 | 0.995 |
|  | CTMA-1473 | 68 365 | 71 - 47 336 | 4593 | 0.996 |
|  | CTMA-1402 | 74 836 | 80 - 54 138 | 2450 | 0.995 |
|  | CTMA-1421 | 63 781 | 70 - 78 397 | 3095 | 0.999 |
|  | CTMA-1424 | 829 644 | 70 - 138 439 | 639 | 0.999 |

While having lower quality than Illumina, both ONT platforms produced reads that were much longer (Table 3).

As expected, assembled genomes generated by SPAdes with Illumina MiSeq reads were more fragmented (ranging from 74 to 89 contigs) than those produced by canu using MinION and GridION reads (from 2 to 5 contigs). As shown in Table 4, MLVA profiles were generated using the *MLVAType* algorithm on WGS data from nine previously reported isolates (1, 4). The results were perfectly concordant, regardless of the sequencing platforms.

**Table 4.** MLVA profiles of *V. cholerae* isolates obtained from WGS data using the *MLVAtype* shiny application

| Isolate | Miseq-derived<br>MLVA profile | MinION-derived<br>MLVA profile | GridION-derived<br>MLVA profile |
| --- | --- | --- | --- |
| CTMA-1402 | (9,7,7,10,16) | Idem | Idem |
| CTMA-1421 | (9,7,7,11,17) | Idem | Idem |
| CTMA-1424 | (10,7,7,11,15) | Idem | Idem |
| CTMA-1426 | (10,7,6,26,20) | Idem | Idem |
| CTMA-1427 | (10,7,6,24,21) | Idem | Idem |
| CTMA-1432 | (10,7,6,16,20) | Idem | Idem |
| CTMA-1435 | (10,7,6,23,18) | Idem | Idem |
| CTMA-1461 | (11,7,7,13,16) | Idem | Idem |
| CTMA-1473 | (10,7,7,12,16) | Idem | Idem |

The phylogenomic trees derived from MiSeq and ONT genomes are illustrated in Figure 1. As expected, the Illumina-based phylogenomic tree outperformed its ONT-based counterpart in terms of resolution. The difference is due to ONT’s lower sequencing accuracy, which causes increased noise and makes it difficult to identify and characterise SNPs position accurately.

**Figure 1.**
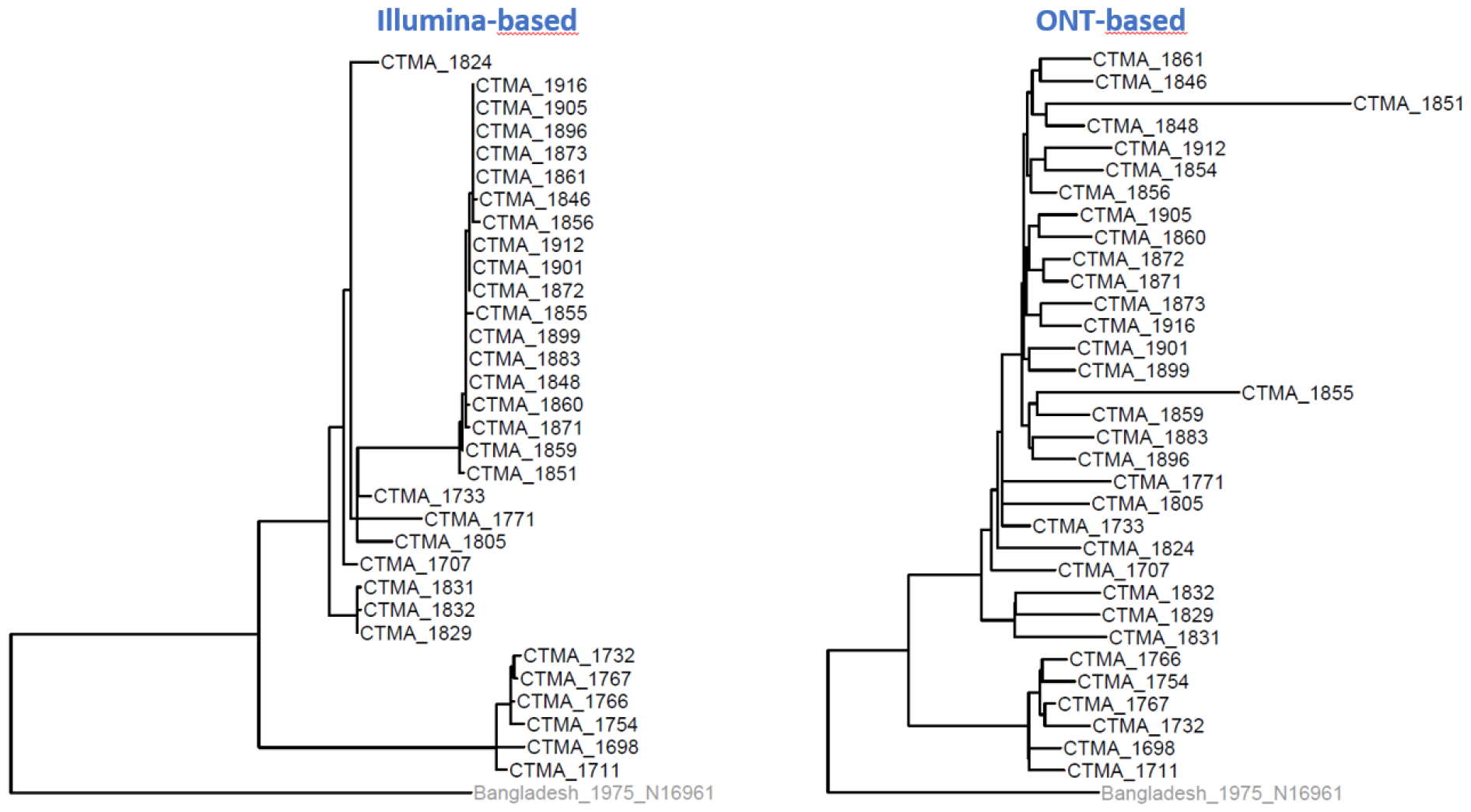
Phylogenomic tree derived from Illumina short-read vs. ONT long-read genome assemblies using kSNP.

The SNP-based phylogenomic tree constructed from Illumina data was compared to MLVA profiles derived from ONT data (Figure 2). Notably, identical MLVA profiles clustered together in the phylogenomic analysis, demonstrating remarkable agreement between both independent approaches.

**Figure 2.**
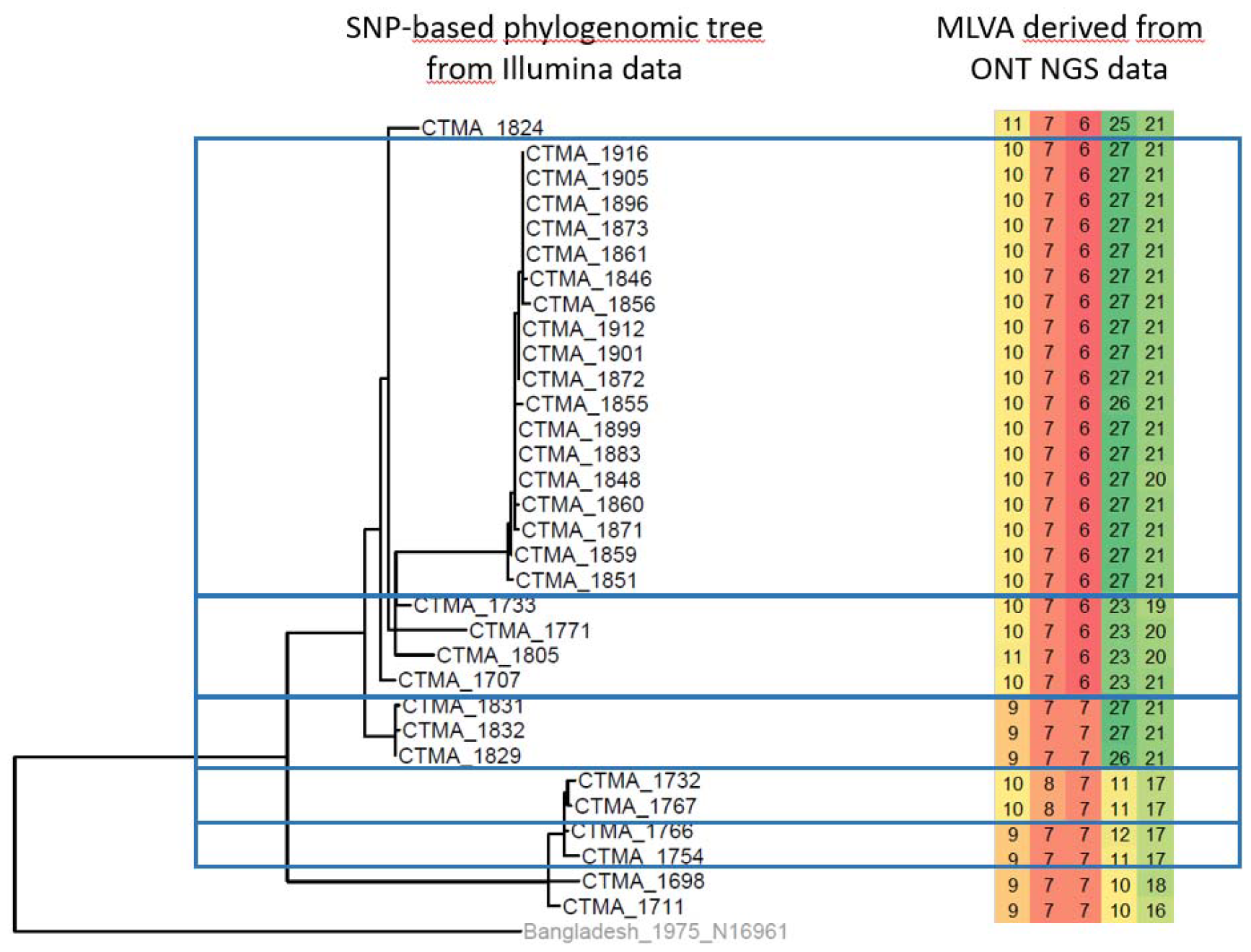
Analysis of MLVA profiles within clusters of isolates defined by an Illumina SNP-based phylogenomic analysis .

## Discussion

Due to its low cost and rapid turnaround time, ONT sequencing MinION and GridION are attractive sequencing platforms for clinical laboratories with a clear potential to replace traditional typing methods. However, this type of analysis is not yet affordable in all institutions because it introduces many new challenges (e.g. data storage, computing power, bioinformatics expertise). Moreover, sequencing with ONT platforms still faces the problem of base-calling accuracy compared to other sequencing platforms such as Illumina short-reads sequencer (8). Accordingly, the present study was designed to assess the impact of the lower sequencing accuracy of the ONT technology on assembled genomic region characterized by variable number of tandem repeats, using the MinION and GridION sequencing platforms. Considering that we previously demonstrated the accuracy of Illumina MiSeq-derived MLVA profiles on *V. cholerae* isolates, we compared the ONT results with the Illumina results taken as a reference. In the first phase of the study, we used the same *V. cholerae* isolates as previously (1, 4) and assembled the contigs with SPAdes.

The MLVA profiles of two isolates (CTMA-1424 and CTMA-1426) differed between our current and previous studies, highlighting the potential instability of MLVA profiles over time in cultured isolates. This phenomenon was previously reported by Kendall et. al. (9) for *V. parahaemolyticus*, where it was observed after 30 passages. It is noteworthy that, in our case, fewer than three passages occurred between the start of the previous study and the end of the current study.

In the current study, the perfect concordance of the results generated by the three sequencing platforms demonstrates the backward compatibility of ONT WGS sequencing with conventional MLVA typing. It is noteworthy that, in this study, MiSeq-derived MLVA profiles were free of censored values due to two factors: (1) the limited number of repetitions per motif (maximum of 26 for the 4th motif of CTMA-1426) and (2) the use of a long k-mer size (175) during genome assembly with SPAdes v.3.13.0.

In the second phase of the study, we compared phylogenetic inferences derived from short-read (Illumina) and long-read (ONT) sequencing technologies. This analysis revealed that Illumina short-read sequencing provides better resolution for phylogenomic reconstruction than ONT-based approaches. Notably, there is a striking agreement between groups of genetically similar isolates, referred to as ‘isolate clusters’, identified using ONT-based MLVA profiles and those derived from the short-read-based phylogenomic analysis.

## Data Availability

All NGS data are available from the European Nucleotide Archive (ENA, http://www.ebi.ac.uk/ena), available under study accession number PRJEB55717

## Ethical standards

Not applicable

## References

1. Ambroise J, Irenge LM, Durant JF, Bearzatto B, Bwire G, Stine OC, et al. Backward compatibility of whole genome sequencing data with MLVA typing using a new MLVAtype shiny application for Vibrio cholerae. PLoS One. 2019;14(12):e0225848.

2. Bankevich A, Nurk S, Antipov D, Gurevich AA, Dvorkin M, Kulikov AS, et al. SPAdes: a new genome assembly algorithm and its applications to single-cell sequencing. J Comput Biol. 2012;19(5):455–77.

3. Delahaye C, Nicolas J. Sequencing DNA with nanopores: Troubles and biases. PLoS One. 2021;16(10):e0257521.

4. Irenge LM, Ambroise J, Mitangala PN, Bearzatto B, Kabangwa RKS, Durant JF, et al. Genomic analysis of pathogenic isolates of Vibrio cholerae from eastern Democratic Republic of the Congo (2014-2017). PLoS Negl Trop Dis. 2020;14(4):e0007642.

5. Irenge LM, Ambroise J, Bearzatto B, Durant JF, Wimba LK, Gala JL. Genomic evolution and rearrangement of CTX–Φ prophage elements in Vibrio cholerae during the 2018–2022 cholera outbreaks in The Democratic Republic of Congo. medRxiv. 2024.

6. Koren S, Walenz BP, Berlin K, Miller JR, Bergman NH, Phillippy AM. Canu: scalable and accurate long-read assembly via adaptive k-mer weighting and repeat separation. Genome Res. 2017;27(5):722–36.

7. Gardner SN, Hall BG. When whole-genome alignments just wonxg’t work: kSNP v2 software for alignment-free SNP discovery and phylogenetics of hundreds of microbial genomes. PLoS One. 2013;8(12):e81760.

8. Petersen LM, Martin IW, Moschetti WE, Kershaw CM, Tsongalis GJ. Third-Generation Sequencing in the Clinical Laboratory: Exploring the Advantages and Challenges of Nanopore Sequencing. J Clin Microbiol. 2019;58(1).

9. Kendall EA, Chowdhury F, Begum Y, Khan AI, Li S, Thierer JH, et al. Relatedness of Vibrio cholerae O1/O139 isolates from patients and their household contacts, determined by multilocus variable-number tandem-repeat analysis. J Bacteriol. 2010;192(17):4367–76.

